# Spectrum of spinal cord involvement in COVID-19: A systematic review

**DOI:** 10.1101/2020.09.29.20203554

**Authors:** Ritwick Mondal, Shramana Deb, Gourav Shome, Upasana Ganguly, Durjoy Lahiri, Julian Benito Leon

## Abstract

**Background and aims:** Recent reports reveal incidences of spinal cord involvement in form of para-infectious or post-infectious myelitis raising potential concerns about the possibilities of SARS-CoV-2 behind the pathogenesis of spinal cord demyelination. In this study, we intend to summarise so far available pieces of evidence documenting SARS-CoV-2 mediated spinal demyelination in terms of clinical, laboratory parameters and imaging characteristics.

**Methodology:** This review was carried out based on the existing PRISMA (Preferred Report for Systemic Review and Meta-analyses) consensus statement. Data was collected from four databases: Pubmed/Medline, NIH Litcovid, Embase and Cochrane library and Preprint servers up till 10^th^ September, 2020. Search strategy comprised of a range of keywords from relevant medical subject headings which includes “SARS-COV-2”, “COVID-19”, “demyelination” etc.

**Results:** A total of 21 cases were included from 21 case reports after screening from various databases and preprint servers. Biochemical analysis reveals that the majority of cases showed elevated CSF protein as well as lymphocytic pleocytosis. Interestingly, a majority of cases were found to be associated with long extensive transverse myelitis (LETM), and remaining cases were found to be associated with isolated patchy involvement or isolated short segment involvement or combined LETM and patchy involvement. Few cases were also found with significant co-involvement of the brain and spine based on the imaging data.

**Conclusion:** It can be interpreted that SARS-CoV-2 may play a potential role in spinal demyelinating disorders in both para-infectious and post-infectious forms.

**Highlights:**

- Imaging data reveals LETM, short and patchy involvements
- Para infectious myelitis precedes post-infectious manifestation
- Altered CSF parameters and myelitis-like symptoms at the onset of COVID-19
- Similar spinal cord involvements in related HCoVs infections

## 1 Introduction

COVID-19 pandemic has now wreaked havoc across the globe for around 6 months since WHO declared it as a pandemic on March 11, 2020. Initially, it was considered to be primarily a respiratory pathogen. However, with time it has been understood as a virus with the potential to cause multi-system involvement. Among reports of various organ involvements in COVID-19 neurological manifestations have drawn significant attention [1]. It is now known that diverse central and peripheral nervous system features may appear following SARS-CoV-2 infection [2]. Neurological features may occasionally precede the typical constitutional or respiratory symptoms of COVID-19 [3]. Among the CNS features stroke is the most frequently reported manifestation while demyelination [4] and seizures [5] are also being increasingly documented. COVID-19 related PNS involvement has mostly been seen in the form of Guillain-Barre Syndrome (GBS) and myositis [6].

In recent times, several reports have come up describing spinal cord involvement in the context of COVID-19. Ranging from vascular to demyelinating pathology, various forms of spinal cord involvement have been documented. Additionally, there are reports revealing cord enhancement in association with polyradiculoneuropathy [4]. In sum, spinal cord involvement in COVID-19 has been steadily receiving attention for some time now [7].

Seemingly the inter-relationship between demyelinating disorders and COVID-19 has two facets. SARS-CoV-2 infection may lead to spinal cord as well as brain demyelination. On the other hand, patients with known primary demyelinating disorders may experience an exacerbation of pre-existing neurological features. In the present systematic review, we set out with the objective of organizing the so far available evidence of spinal cord demyelination in the setting COVID-19 in terms of clinical presentation, laboratory features, and imaging characteristics. The presented summary of COVID-19 associated myelitis has important prognostic as well as therapeutic implications from the perspective of clinical neurology.

## 2 Methodology

### 2.1 Design

This systematic review was conducted by following the Preferred Reporting for Systematic Review and Meta-Analysis (PRISMA) consensus statement (CRD42020201843) [https://www.crd.york.ac.uk/prospero/display_record.php?ID=CRD42020201843]. Studies relevant to the confirmed cases of COVID-19 infection with a confirmed or suspected association of demyelinating disorders of the spinal cord were included.

### 2.2 Search strategy

In this systemic review four databases: Pubmed/Medline, NIH LitCovid, Embase, and Cochrane Library were searched using pre-specified searching strategies, and this search was concluded on September 10, 2020. The search strategy consists of a variation of keywords of relevant medical subject headings (MeSH) and keywords, including “SARS-CoV-2”, “COVID-19”, “coronavirus”, “demyelinating disorders”, “Multiple sclerosis” and “Encephalomyelitis”. Severe Acute Respiratory Syndrome Coronavirus (SARS-CoV) and Middle East Respiratory Syndrome Coronavirus (MERS-CoV) were also included in our search strategy to capture related articles. We also hand-searched additional COVID-19 specific articles using the reference list of the selected studies, relevant journal websites, and renowned pre-print servers (medRxiv, bioRxiv, pre-preints.org) from 2019 to the current date for literature inclusion. To decrease publication bias, we invigilated the references of all studies potentially missed in the electrical search. Content experts also searched the grey literature of any relevant articles.

### 2.3 Study selection criteria

All peer-reviewed, pre-print (not-peer-reviewed) including cohort, case-control studies, and case reports which met the pre-specified inclusion and exclusion criteria were included in this study.

### 2.4 Inclusion criteria

Studies meeting the following inclusion criteria were included : (i) Conducted for the COVID-19 positive patients with suspected or confirmed demyelinating disorders of spinal cord. (ii) Studies revealing possible association of multiple sclerosis (MS) or related neuroautoimmune disorders with confirmed or suspected spinal involvement in COVID-19 .(iii) Simultaneously parallel search was conducted to have a comparative as well as a retrospective outlook into the distribution and incidences of similar neurological manifestations in previous outbreak i.e. Severe Acute Respiratory Syndrome (SARS-CoV) and Middle East Respiratory Syndrome Coronavirus (MERS-CoV) and various other HCoVs. (iv) Studies published in the English language were included for qualitative synthesis in a narrative review.

### 2.5 Exclusion criteria

Studies excluded if COVID-19 was not confirmed among patients and written in languages other than English. We also excluded review papers, viewpoints, commentaries, and studies where no information related to neurological manifestations or spinal demyelination was reported.

### 2.6 Data Extraction

Before the screening process, a team of two reviewers (GS and SD) participated in calibration and screening exercises. The first reviewer (GS) subsequently screened independently the titles and abstracts of all identified citations, and the second reviewer (SD) verified those citations and screened papers by (GS). Another reviewer (UG) then retrieved and screened independently the full texts of all citations deemed eligible by the reviewer (SD) and analyzed those data. The other two reviewers (RM and DL) independently verified these extracted full texts for eligibility towards analysis and designed the overall study structure. The corresponding author (JBL) had resolved disagreements whenever necessary and took final decisions regarding the study. Throughout the screening and data extraction process, the reviewers used piloted forms. In addition to the relevant clinical data, the reviewers also extracted data on the following characteristics: study characteristics (i.e. study identifier, study design, setting, timeframe); outcomes (qualitative and/or quantitative); clinical factors (definition and measurement methods); study limitations. The Newcastle-Ottawa scale was used to assess the selection procedure, the comparability, and the outcomes of each reviewed study.

### 2.7 Statistical analysis

Both qualitative and quantitative data were expressed in percentages. Unit discordance among the variables was resolved by converting the variables to a standard unit of measurement. A value of ‘p’<0.05 was considered as statistically significant but it could not be calculated due to insufficient data. A meta-analysis was planned to analyze the association of the demographic findings, symptoms, biochemical parameters, outcomes but was later omitted due to lack of sufficient data.

## 3 Result

Around 750 articles were initially identified from the aforementioned databases and 253 articles were identified from different pre-prints servers. Finally, a total of 712 articles were identified from different databases searched after removing the duplicates. Of the selected articles, 600 articles were excluded after screening titles and abstracts, leaving 112 articles for full-text review for possible inclusion in this study. Of these, 60 articles were excluded based on the inclusion and exclusion criteria for the study sample (e.g., excluded demyelination of only brain, pre-existing demyelination related articles), and few other articles were excluded for study types (e.g review papers, correspondence, viewpoints, commentaries). A total of 52 articles were finally selected for this study; 21 articles were included in the analysis, and the remaining 31 articles were synthesized narratively [**Figure 1**].

**Figure-1.**
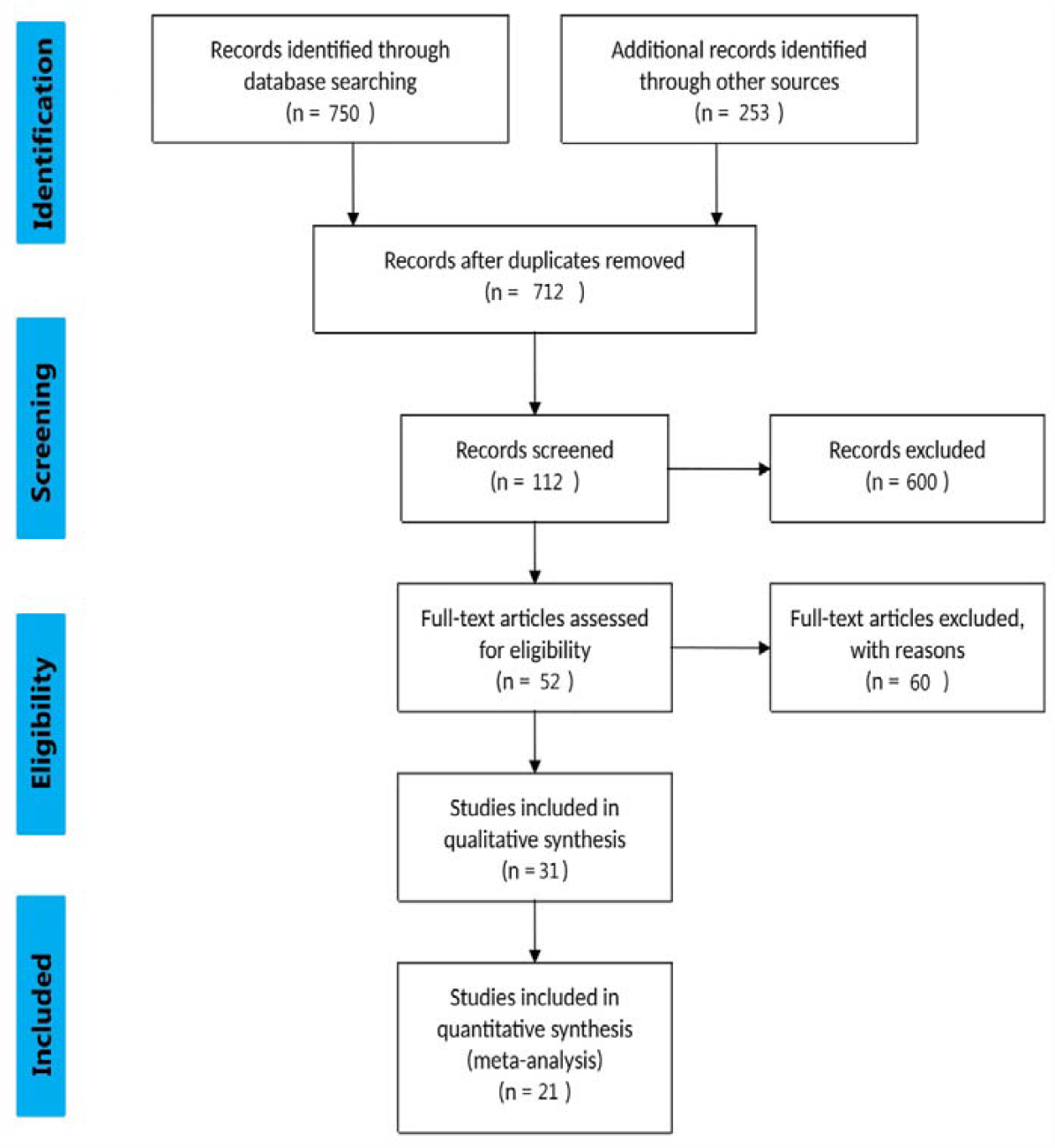
Flow chart showing the algorithm used to identify the studies based on spinal involvement in COVID-19 that met inclusion criteria. Flow diagram template adopted from PRISMA

The selected study comprises of 21 case reports with 52% (n=11) male and 48% (n=10) female along with the mean age around (46.66±17.97). They had developed various COVID-19 related symptoms such as fever (n=11, 52%),cough (n=6, 29%), myalgia (n=6, 29%),vomiting (n=2, 10%),nausea (n=1, 5%)and many more. The neurological symptoms reported were weakness (n=14, 66.66%), sensory deficit (n=14, 66.66%) ataxia (n=1, 4.76%), autonomic dysfunction (n=8, 38.09%),cognitive impairment (n=2, 9.52%).

Various CSF parameters under biochemical analysis such as Glucose (mg/dl) (n=5, 24%), Protein (mg/dl) (n=13, 70%), Adenosine Deaminase (μ/L) (n=1, 5%), Lactate Dehydrogenase (unit/L) (n=1, 5%) were recorded with significant levels of elevation from the normal range. Moreover certain cell count parameters such as WBC (n=2, 10%), RBC (n=2, 10%) was significantly higher than the normal range. Cases with Lymphocytic pleocytosis (n=5, 24%) were also reported. SARS-COV-2 was detected in (n=5, 24%) cases using RT-PCR (n=4) and IgG positive (n=1). SARS-COV-2 detection came negative in (n=13, 62%) cases whereas SARS-COV-2 testing was not done in two cases and data was unavailable for one case. Autoimmune profiling for majority of cases came negative except two cases with positive oligoclonal band (OCB) and lupus antigen respectively [**Table-1**].

**TABLE-1.** Demographic and clinical features.

| Serial no. | Authors | Sex (M/F)<br>Age (yrs) | Co-morbidities | Neurological signs/symptoms | CSF characteristics |  | Clinical diagnosis proposed by authors | Clinical features of Covid19 infection | Latency (no. of days) | Treatment | Outcome |
| --- | --- | --- | --- | --- | --- | --- | --- | --- | --- | --- | --- |
|  |  |  |  |  | SARS-Cov2 detection | CSF parameters |  |  |  |  |  |
| 1 | Abdelhady *et al* [Qatar] | M, 52 | T2DM, G6PD deficiency | Lower abdominal pain and inability to pass urine. Bilateral lower limb weakness (flaccid paralysis). No sensory affection. | Negative | Elevated WBCs (particularly lymphocytes) and elevated proteins | Acute flaccid myelitis | Fever | NA | Steroids and Acyclovir | Death due to cardiac arrest |
| 2 | Alketbi *et al* [UAE] | M, 32 | NA | Bilateral lower limb weakness, difficulty in sitting up, passing urine. Sensory deficit, weakness involved in the whole lower limbs. Hypotonic muscle tone in both lower limbs. Reflexes 2+ in upper and 1+ in lower limbs, no association of bulbar symptoms | NA | NA | Acute myelitis | Fever with flu-like symptoms | 1 | Methylprednisolone (1g/d for 5 days), Acyclovir, Enoxaparin | Still in treatment |
| 3 | Chakraborty U. *et al* [India] | F, 59 | Obesity | Acute, progressive, ascending flaccid paraplegia of both lower limbs (MRC 0/5) along with urinary retention and constipation. Hypotonia in both lower limbs. Deep tendon reflexes of both lower limbs were absent with bilateral mute plantar response. No sensation below T10 segment level. profound hypotonia of both lower limbs. | Negative | Elevated protein (71.4 mg/dl) and adenosine deaminase (4.5µ/L) Glucose (75mg/dl) | Acute transverse myelitis | Fever | NA | Methylprednisolone (1g/d) with antipyretics and supportive care | Death due to cardiac arrest |
| 4 | Chow *et al* [Australia] | M, 60 | Well controlled hypertension and hypercholesterolaemia | Bilateral lower limb weakness with constipation, urinary retention, hyperreflexia, reduced proprioception of lower limbs, patchy paresthesia to the level of umbilicus. | Negative | Elevated protein (79 mg/dl) Glucose (57.66mg/dl) Lactate dehydrogenase (<30 unit/L) | Acute transverse myelitis | Fever, dysgeusia, anosmia, cough | 2 | Methylprednisolone (1g/d for 3 days) | Discharged |
| 5 | Domingues *et al* [Brazil] | F, 42 | Neurological symptoms | Paresthesia of upper left limb progressing to left hemithorax and hemiface | Positive | Protein 32 mg/dl, glucose 68 mg/dl OCB (negative) | Clinically isolated syndrome (CIS) and suspected spinal demyelination | Mild respiratory symptoms including coryza and nasal obstruction | NA | NA | Discharged |

TABLE-1 (continuation)
| Serial No. | Authors | Sex (M/F)<br>Age (yrs) | Co-morbidities | Neurological signs/symptoms | CSF characteristics |  | Clinical diagnosis proposed by authors | Clinical features of Covid19 infection | Latency (no. of days) | Treatment | Outcome |
| --- | --- | --- | --- | --- | --- | --- | --- | --- | --- | --- | --- |
|  |  |  |  |  | SARS-Cov2 detection | CSF parameters |  |  |  |  |  |
| 6 | Kaur *et al* [USA] | F, 3 | NA | Progressive extremity weakness and decreased sensation. Progressed to flaccid quadriparesis and neurogenic respiratory failure requiring intubation. Complete quadriplegia after 12 hours. | Negative | Elevated RBC (282/mm<sup>3</sup>), total nucleated cells (42/mm<sup>3</sup>), neutrophil (96%), protein (58mg/dl) | LETM | No symptoms | 21 | Methylprednisolone (30mg/kg/d) for 5 days, IVIG (2g/kg for 5 days), seven sessions of plasma exchange (1.0-1.5 volumes exchange per session). rituximab 4 infusions 375 mg/m<sup>2</sup> | Discharged |
| 7 | Lisnic *et al* [Moldova] | M, 27 | HIV infection for 1 year, on ART | Paresthesia and numbness in legs and right arm, paralysis in lower extremities, bladder and bowel dysfunction, spastic tetraparesis with 4/4,5 MRC in upper and 0.5/2 MRC in lower extremities. Th7 superficial and C7 deep reflexes.sensory level disturbances | Negative | Profiling normal OCB (negative) | Acute transverse myelitis | Subfebrile (mild fever) | NA | Methylprednisolone (1g/d for 5 days), five plasma exchange sessions | Still in treatment |
| 8 | Maideniuc *et al* [USA] | F, 61 | Hypertension,hyperlipidemia, hypothyroidism, remote history of nasopharyngeal and uterine cancer | Tingling sensation in fingers and toes. Lost sensation from chest down and progressive weakness in extremities. Bowel and bladder retention. Brisk reflexes and increased tone in lower extremities. Sensory level at C3. | Negative | Increased RBC (312/mm<sup>3</sup>), protein (87mg/dl), glucose (73mg/dl) OCB (negative) IgG index normal | Acute necrotizing myelitis with AMAN | Runny nose and chills | 4 | Methylprednisolone (1g/d for 5 days), five plasma exchange sessions | Discharged |
| 9 | Mc Cuddy *et al* [USA] | F,40 (patient 1) | T2DM,HTN, obesity, and pregnancy 30 weeks gestation | Diffuse weakness noted post extubation,plegic in legs bilaterally,symmetric weakness in upper extremity,DTRs brisk,sensation intact | Negative | Protein-95 mg/dl Glucose-85 mg/dl Cell count – WBC 2/uL OCB (negative) | ADEM | Cough,chest pain,fever,shortness of breath | NA | Decadron 20 mgIV (for 5 days) and 10 mg (for 5 days) Hydroxychloroquine , Zinc,convalescent plasma therapy | Improving.Partial return of strength in upper extremities core. Regaining some function in distal lower extremities. Cognition intact. |

TABLE-1 (Continuation)
| Serial No. | Authors | Sex (M/F)<br>Age (yrs) | Co-morbidities | Neurological signs/symptoms | CSF characteristics |  | Clinical diagnosis proposed by authors | Clinical features of Covid19 infection | Latency (no. of days) | Treatment | Outcome |
| --- | --- | --- | --- | --- | --- | --- | --- | --- | --- | --- | --- |
|  |  |  |  |  | SARS-Cov2 detection | CSF parameters |  |  |  |  |  |
| 10 | Munz *et al*<br>[Germany] | M, 60 | Hypertension, mild fatty liver, ureterolithiasis | Bladder dysfunction. Progressive weakness of lower limbs. Hypesthesia below Th9 level and moderate spastic paraparesis. Babinski sign positive bilaterally. Cognition and cranial nerves unaffected. | Negative | Abnormal lymphocytic pleocytosis (16/μl), protein (79.3 mg/dl) OCB (negative) | Acute transverse myelitis | Typical respiratory features | 2 | Acyclovir, ceftriaxone, methylprednisolone (100mg/d) | Discharged with steroid taper scheme |
| 11 | Novi *et al*<br>[Italy] | F, 64 | Vitiligo, hypertension, monoclonal gammopathy | Irritability, headache, bilateral pupillary defect, visual loss, right abdominal sensory level deficit, left sided lower limb hyperreflexia with Babinski sign | Positive | Lymphocytic pleocytosis (22 cell/mm<sup>3</sup>) with mild hyperproteinorria (45.2 mg/dl) OCB-(positive) | Suspected ADEM | Flu-like symptoms<br>Anosmia, ageusia | 25 | Methylprednisolone (1g/d for 5 days), oral prednisolone (75mg/d), IVIG (2g/kg for 5 days) | Discharged with oral prednisolone tapering |
| 12 | Otluglu *et al*<br>[Turkey] | M, 48 | None | Progressive headache. Normal neurological examination. | Positive | Protein 40mg/dl<br>Glucose- 90.09 mg/dl<br>No cell detected microscopically | Covid-19 pneumonia with viral encephalitis and myelitis | Persistent cough, fatigue, myalgia, anosmia | NA | HCQ, Favipiravir, Acyclovir.(3*700 mg IV)<br>Methylprednisolone (1g/d for 5 days).<br>Levitiracetam, Piperacillin (3*4.5 g IV), Tazobactam (3*4.5 g IV), | Still under treatment with stable condition |
| 13 | Sarma *et al*<br>[USA] | F, 28 | Hypothyroidism | Persisting lumbosacral back pain without radiation. Paresthesias in lower extremities with total loss of sensation upwards till tip of tongue. Urinary retention. Decreased sensation below 5<sup>th</sup> thoracic vertebra, proprioception, positive Lhermitte's sign | NAD | Elevated mononuclear cells (125/μl), protein (60mg/dl) Normal level glucose | Transverse myelitis | Cough, low grade fever, low back pain, myalgia, rhinorrhea, nausea and vomiting. | NA | Prednisolone and two plasma exchange sessions | Discharged with steroid tapering |

TABLE-1 (Continuation)
| Serial No. | Authors | Sex (M/F)<br>Age (yrs) | Co-morbidities | Neurological signs/symptoms | CSF characteristics |  | Clinical diagnosis proposed by authors | Clinical features of Covid19 infection | Latency (no. of days) | Treatment | Outcome |
| --- | --- | --- | --- | --- | --- | --- | --- | --- | --- | --- | --- |
|  |  |  |  |  | SARS-Cov2 detection | CSF parameters |  |  |  |  |  |
| 14 | Sotoca *et al* [Spain] | F, 69 | NA | Irradiated cervical pain, imbalance and motor weakness. Numbness in left hand. Right facial and left hand hypesthesia, left hand weakness, general hyperreflexia | Negative | Lymphocytic pleocytosis (75 cells/microleter) and hyperproteinorraquia (28.3mg/dl) OCB negative | Acute necrotizing myelitis | Fever, dry cough | 1 | Methylprednisolone (1g/d for 5 days) Plasma exchange | Discharged with oral prednisolone |
| 15 | Utukuri *et al* [USA] | M, 44 | NA | urinary retention, bilateral lower extremity weakness, numbness, inability to walk | Initially positive, later negative | WBC 0.6/microleter lymphocytes 92% Protein- 26mg/dl OCB-negative | ADEM | Lethargy | NA | Methylprednisolone and IVIG | Discharged |
| 16 | Valiuddin *et al* [USA] | F, 61 | NA | Numbness and tingling in hands and feet progressing to severe weakness in lower extremities bilaterally. Difficulty in standing and ambulation. Numbness ascending to abdomen with constipation. Difficulty in voiding. Bilateral weakness in lower extremity (3/5), upper extremity (4/5). Decreased ankle reflexes. Bilateral extensor plantar responses. | Negative | Elevated protein (87 mg/dl) and albumin (53.5 mg/dl) WBC-1/mm<sup>3</sup> OCB (negative) | Acute transverse myelitis | Generalized weakness, rhinorrhea, chills | 4 | Methylprednisolone and five plasma exchange sessions | Still in treatment |
| 17 | Zachariadis *et al* [Switzerland] | M, 63 | Obesity, | Headache, paresthesia, hypesthesia in feet progressing to abdomen. Moderate paresis of lower limbs with pyramidal signs and sensory level at T10 | Negative | Slight elevation of leukocytes and protein (57.3 mg/dl) WBC 16 /microleter | Transverse myelitis | Rhinorrhea, odynophagia, myalgia, fever | 5 | IVIG (0.4g/kg) for 5 days , corticosteroid for 5 days | Discharged |

TABLE-1 (Continuation)
| Serial No. | Authors | Sex (M/F)<br>Age (yrs) | Co-morbidities | Neurological signs/symptoms | CSF characteristics |  | Clinical diagnosis proposed by authors | Clinical features of Covid19 infection | Latency (no. of days) | Treatment | Outcome |
| --- | --- | --- | --- | --- | --- | --- | --- | --- | --- | --- | --- |
|  |  |  |  |  | SARS-Cov2 detection | CSF parameters |  |  |  |  |  |
| 18 | Zanin *et al* [Italy] | F, 54 | Anterior communicating artery aneurysm 20 years ago, treated surgically | Found unconscious at home. GCS of 12 (E3 M6 V3) without focal sensorimotor deficits, incontinence | Negative | Normal | Spinal demyelinating lesion | Anosmia and Ageusia. | NA | Dexamethasone (20mg/day for 10 days then 10mg/day for 10 days). Lacosamide, levetiracetam, phenytoin Antiretroviral and hydroxychloroquine, antiepileptic therapy (phenytoin, levetiracetam, lacosamide) | Discharged |
| 19 | Zhao *et al* [China] | M, 66 | NA | Weakness in both lower limbs with urinary and bowel incontinence Hyporeflexia (0/5 power reduction) in bilateral lower extremities, sensation impaired in both legs. Sensory level at T10 to pin-prick testing with feelings of parasthesia and numbness below the level.<br>Decreased tendon reflexes of both lower limbs. Pain, temperature and tactile sensations lowered below chest level 10.. | ND | ND | Acute myelitis | Fever, fatigue | 6 | Lopinavir/Ritonavir, ganciclovir. Dexamethasone (10mg/d), IVIG (15g/d), moxifloxacin, meropenem, glutathione, mecobalamin, pantoprazole | Discharged |
| 20 | Zhou *et al* [USA] | M, 26 | NA | Bowel and bladder dysfunction, cognitive and mood changes, numbness on lower extremities, neck discomfort with forward flexion, electric like pain, sequential vision loss (left then right) | Negative | Protein -31mg/dl<br>Glucose-57mg/dl<br>Cell count 55/microleter (100% WBC) | MOG-Ab associated optic neuritis with myelitis | No significant associations | NA | Methylprednisolone (IV) followed by prednisolone taper | Complete resolution and subsequent follow up |

**TABLE-1 (Continuation)**
| Serial No. | Authors | Sex (M/F)<br>Age (yrs) | Co-morbidities | Neurological signs/symptoms | CSF characteristics |  | Clinical diagnosis proposed by authors | Clinical features of Covid19 infection | Latency (no. of days) | Treatment | Outcome |
| --- | --- | --- | --- | --- | --- | --- | --- | --- | --- | --- | --- |
|  |  |  |  |  | SARS-Cov2 detection | CSF parameters |  |  |  |  |  |
| 21 | Zoghi *et al*<br>[Iran] | M, 21 | NA | Deep tendon reflexes in all 4 limbs. No Babinski sign. Impaired position and light touch sensation in lower limbs. T8 sensory level. Absence of abdominal cutaneous reflex,urinary retention,paresthesia,weakness in upper limbs | IgG positive | high protein (281 mg/dl)<br>glucose (34 mg/dl) | LETM | Fever, cough, sore throat, loss of appetite, vomiting, malaise | 17 | Plasma exchange (250ml for 5 days), intravenous vancomycin, meropenem, acyclovir | Discharged |

\* LETM –Long extensive transverse myelitis
\*ADEM-Acute disseminated encephalomyelitis
\*T2DM-Type 2 diabetes mellitus
\*HTN-Hypertension
\*AMAN-Acute motor axonal neuropathy
\*DTR-Deep tendon reflex
\*GCS-Glasgow coma scale
\*G6PD-Glucose-6-phosphate dehydrogenase
\*HIV-Human immuno deficiency virus
\*ART-Antero retroviral therapy
\*OCB-Oligoclonal band
\*MOG-Ab-Myelin oligodendrocyte glycoprotein antibody
\*MRC- Medical Research Council
\*IVIG-Intravenous immune globulin

Neuroimaging findings were reported in 20 studies consisting of 20 cases and the remaining 1 case imaging data were not available. Among the imaging modalities, MRI of the spine was done for all the 18 cases whereas brain MRI was found for 16 cases. CT scan of the spine and brain were performed for 1 and 2 cases respectively. Interestingly, we found a majority of cases (n=9, 45%) were associated with isolated LETM whereas a combination of both LETM and patchy was 10% (n=2). Imaging data also reveals that 10% (n=2) cases were associated with isolated patchy involvement and 25% cases (n=5) were associated with isolated short segment involvement. Furthermore, we found co-involvement of both brain and spine for 30% (n=6) cases [**Table-2 and Figure-2**].

**TABLE-2.** Imaging features of included cases.

| SL.no | Reference | Spinal Involvement |  |  | Level of involvement | Brain involvement | Autoimmune profiling |
| --- | --- | --- | --- | --- | --- | --- | --- |
|  |  | LETM | Patchy | Short |  |  |  |
| 1. | Abdelhady *et al* | + | - | - | T2W1 hyperdensity in ventral horns of grey matter in upper and mid thoracic spinal cord | NAD | ANCA (-ve)<br>ANA (-ve) |
| 2. | Alketbi *et al* | - | + | - | Axial T2W1 of cervical and dorsal spine showing central hyperintense signal of multiple levels. | ND | Anti-LA-(-ve)<br>ANCA -(-ve)<br>RF -(-ve)<br>Anti cardiolipin -(-ve)<br>Anti –beta gp -(-ve) |
| 3. | Chakraborty U *et al* | - | - | + | T2W imaging shows dorsal spine hyperintensity at T<sub>6</sub>– T<sub>7</sub> | ND | ND |
| 4. | Chow *et al* | + | - | - | T2 signal elevation centrally in spinal cord from T<sub>7</sub>.T<sub>10</sub> | NAD | Anti myelin associated gp IgM (-ve)<br>Anti-MOG (-ve)<br>Anti-NMO IgG (-ve) |
| 5. | Domingues *et al* | - | - | + | Sagittal T2 /STIR hyper signal in cervical spine indicating small left lateral ventral lesion with mass effect (0.4 cm) [with Gd contrast] | NAD | Anti SSA (-ve)<br>Anti SSB (-ve) |
| 6. | Kaur *et al* | + | - | - | Swelling of cervical spinal cord with T2W1 hyperintense edema from lower medulla to mid thoracic level | NAD | Negative Rheumatoid evaluation<br>Anti-AQP-4 (-ve)<br>Anti-MOG (-ve) |
| 7. | Lisnic *et al* | + | - | - | Extensive C<sub>4</sub>-T<sub>5</sub> lesion in posterior column on T2W imaging and right lateral column with Gd contrast | N AD | ANA- (-ve)<br>ANCA-(-ve)<br>Anti-AQP-4-(-ve)<br>Anti-MOG-(-ve) |

**TABLE-2 (Continuation)**
| Sl. No | Reference | Spinal Involvement |  |  | Level of involvement | Brain involvement | Autoimmune profiling |
| --- | --- | --- | --- | --- | --- | --- | --- |
|  |  | LETM | Patchy | Short |  |  |  |
| 8. | Maideniuc *et al* | + | - | - | T1W hyperintensity at C<sub>3</sub>-C<sub>4</sub> level | NAD | Autoimmune profiling for<br>Lupus (+ve)<br>Sjogren,MOG,NMOSD and<br>all are negative<br>AQP-4 (-ve) |
| 9. | McCuddy *et al* | - | - | - | NAD | Multiple T2 hyperintense lesions with<br>restricted diffusion involving corpus<br>callosum,bilateral cerebral white matter ,right<br>pons and in the bilateral ventral medulla.no<br>hemorrhagic changes. | Autoimmune profiling of<br>CSF came (-ve) |
| 10. | Munz *et al* | - | - | + | Patchy hyperintensity at Th<sub>9</sub>-Th<sub>10</sub> and Th<sub>3</sub>-Th<sub>5</sub> | NAD | Anti neuronal(-ve)<br>Anti-MOG(-ve)<br>Anti-AQP4(-ve) |
| 11. | Novi *et al* | - | - | + | T2W Sagittal shows hyperintense spindle like single lesion<br>at T<sub>8</sub> level. | Multiple T1 post Gd enhancing lesions in<br>brain.Follow up MRI showed reduction in<br>number of lesions | Anti-AQ4 (-ve)<br>Anti-MOG (-ve) |
| 12. | Otluglu *et al* | - | - | + | Confined hyperintense lesion at upper cervical spine | FLAIR images demonstrate hyperintense<br>signals in posterior medial cortical surface of<br>the temporal lobe | ND |
| 13. | Sarma *et al* | + | - | - | Elongated signal changes throughout the spinal cord to<br>conus medullaris and involving medulla | ND | Anti-ANA (-ve) |
| 14. | Sotoca *et al* | + | - | - | Sagittal T2W shows hyperintensity extending from medulla<br>oblongata to C<sub>7</sub> | NAD | Neuronal surface antibody<br>was ruled out |
| 15. | Utukuri *et al* | + | + | - | T2 hyperintense lesions throughout cervical and thoracic<br>spinal cord with slight cord expansion.<br>Lumbar spine MRI showed hyperintense T2 signal of conus<br>medullaris | Axial FLAIR images demonstrate FLAIR<br>hyperintense lesions within left posterior<br>parietal lobe and periventricular region | Cardiolipin Ab IgM<br>(26agittal26) |

**TABLE-2 (Continuation)**
| SI no. | Reference | Spinal Involvement |  |  | Level of involvement | Brain involvement | Autoimmune profiling |
| --- | --- | --- | --- | --- | --- | --- | --- |
|  |  | LETM | Patchy | Short |  |  |  |
| 16. | Valiuddin *et al* | - | + | - | STIR shows (27 sagittal plane) extensive ill defined patchy hyperintense signal throughout the central aspect of spinal cord. | ND | Anti-GFAP-(-ve)<br>mGLUR (-ve)<br>NMDA-R (-ve)<br>MOG –(-ve) |
| 17. | Zachariadis *et al* | - | - | - | NAD | NAD | Anti ANCA-(-ve)<br>Anti MOG- (-ve)<br>Anti SSB (-ve)<br>Anti SSA (-ve)<br>Rheumatoid factor (-ve)<br>GFAP<br>ANA (-ve)<br>Beta 2 glycoprotein 1 (-ve) |
| 18. | Zanin *et al* | + | + | - | Hyperintense intra-medullary signal alterations in T2 and without contrast enhancement located at bulb-medullary junction at C<sub>2</sub> and from C<sub>3</sub> to Th<sub>6</sub>. | T2W1 hyperintense alterations in periventricular white matter without restriction of diffusion nor contrast enhancement | ND |
| 19. | Zhou *et al* | + | - | - | T2W1 MRI shows spinal signal enhancement at lower cervical and upper Thoracic segment with mild central cord thickening in gd contrast. | T1W1 image bilateral optic nerve thickening (uniform enhancement) upto the peri chiasmal segments | AQP4-(-ve)<br>MOG IgG- (elevated)<br>(1:1000) |
| 20. | Zoghi *et al* | + | - | - | Axial LETM of upper cervical with intramedullary lesion. | Sagittal T2 weighted FLAIRF showed corticospinal tract lesions in internal capsule extending to cerebral peduncles and pons. Heterogenous marble patterned hyperintensity in splenium of corpus callosum without diffusion weighted restrictions or contrast enhancement. | Anti-NMDAR (-ve)<br>MOG-(-ve)<br>AQP4-(-ve)<br>Antiphospholipid (-ve) ,<br>HLAB5 (-ve) ACE (-ve) |

\*T2W1-T2 weighted image \*Gd-gadolinium \*STIR-short T1 inversion recovery \*ANCA-antineutrophil cytoplasmic antibodies \*ANA-antinuclear antibody \*RF-rheumatoid factor \*Anti-SSA- anti Sjogrens's syndrome –related antigen A autoantibodies \*Anti-SSB- anti Sjogrens's syndrome –related antigen B autoantibodies \*AQP-4- aquaporin-4 \*NMO-neuromyelitis optica \*LA-lupus antigen \*FLAIR-fluid attenuated inversion recovery \*GFAP-glia fibrillary acidic protein \*m-GLUR-metabotropic glutamate receptors \*NMDA-R-N-methyl-D-aspartate receptor \*HLA-human leukocyte antigen \*ACE- angiotensin converting enzyme

**Figure-2.**
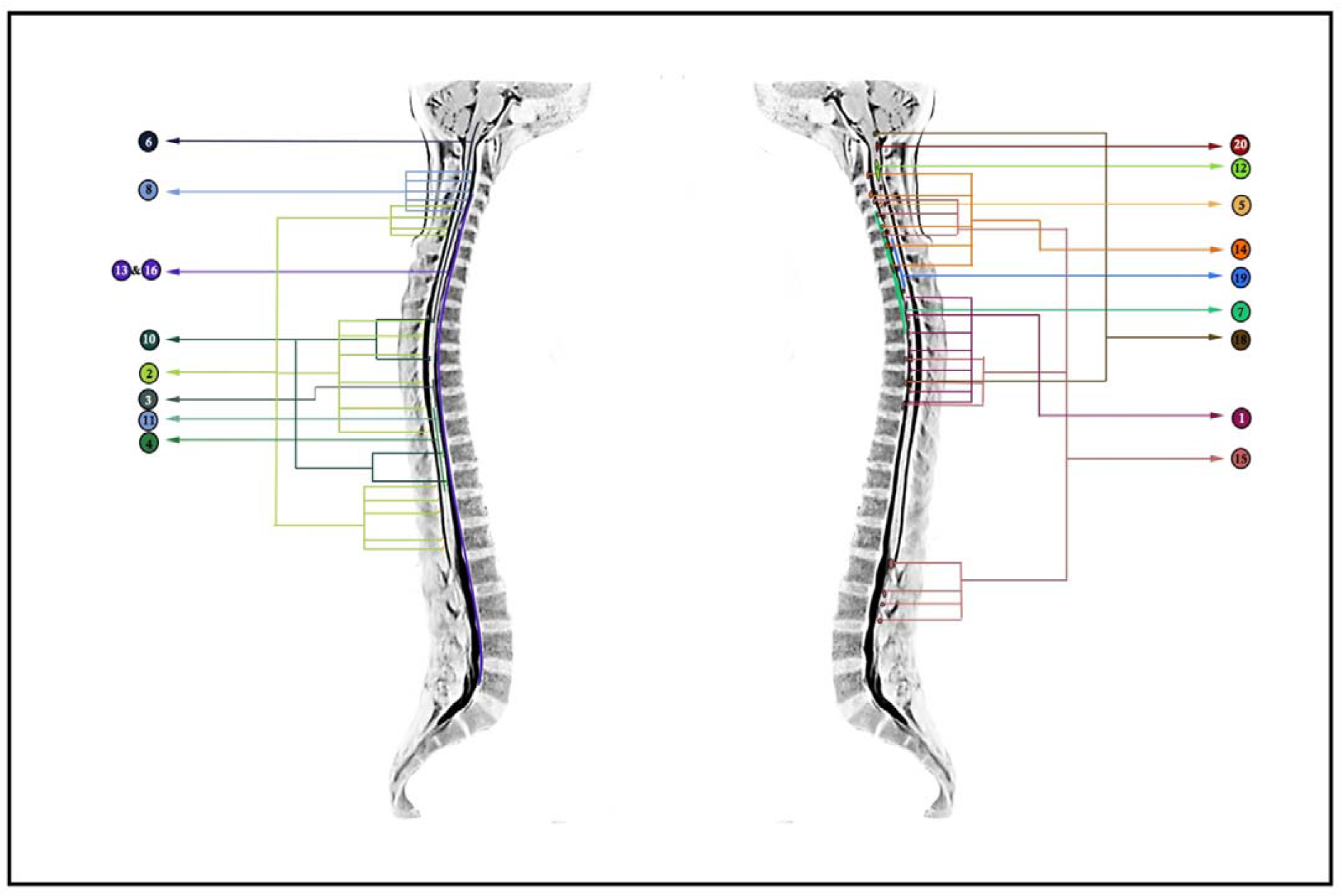
Summarized level and types of spinal cord involvements in COVID-19 as per included case reports (reference number corresponds to Table-2)

Treatment was mainly carried out with corticosteroids (n=18, 86%) like methyprednisolone (n=14, 67%), dexamethasone (n=2, 10%), oral prednisolone (n=1, 5%) etc. Intravenous immunoglobulin was given to (n=5, 24%) cases. Sessions of therapeutic plasma exchange were given to (n=8, 38%) patients. Antivirals drugs such as acyclovir, lopinavir/ritonavir, ganciclovir, favipiravir were administered to (n=7, 33.33%) patients with the most common drug of choice being Acyclovir in (n=5, 24%) cases. Hydroxychloroquine was given to three patients (14.28%). Two cases were reported where anti-epileptic drug was given. A majority of the patients recovered/discharged (n=15, 71%) with mortality of two cases (n=2, 10%) and four patients (n=4, 19%) were still under treatment during the period of study.

## 4 Discussion

During the course of this ongoing pandemic, several immunological features of SARS-CoV-2, affecting different organ systems have been observed by clinicians and researchers across the globe. In the context of neurological manifestations of COVID-19, immunological mechanisms are known to play important roles in giving rise to diverse clinical presentations affecting both CNS as well as PNS [**29**]. Quite antithetical to how SARS-CoV-2 affects PNS (mostly in the form of GBS) [**29**], data regarding spinal cord involvement are scarce. Acute or post-infectious myelitis is not uncommon with several viruses ascribable to its cause, such as Epstein-Barr virus, cytomegalovirus, measles virus, or rhinovirus. In addition, infection-related spinal cord demyelination is known to be associated with brain involvement as well, giving rise to the clinical picture of ADEM.

Spinal cord involvement in COVID-19 might be an under-recognized neurological complication of this novel infectious disease. In the present systematic review, we have found spinal cord involvement in 21 patients with COVID-19 along with their laboratory features and imaging abnormalities. We found 38.1% (n=8) of patients with para-infectious acute myelitis and 14.3% (n=3) with post-infectious acute myelitis. Weakness (66.7%), sensory deficit (66.7%), autonomic dysfunction including sphincter dysfunction (38.1%), and ataxia (4.8%) were the most frequent neurological manifestations at onset.

Besides, neuroimaging data revealed that half of the patients presented with longitudinally extensive transverse myelitis (LETM) followed by patchy (11.1%) and short (27.7%) segment involvement. LETM is usually observed in neuromyelitis optica spectrum disorder (NMOSD), infectious myelitis, lupus-related demyelination, and, occasionally, in MS. In India, tuberculosis is a known cause of LETM [30]. Of interest, half of the reported cases of myelitis in COVID-19 present with long-segment spinal cord involvement. From a clinician’s perspective, this observation basically extends the differential diagnosis of LETM to include COVID-19 related spinal cord demyelination. Due to the small number of available cases, we could not establish a definite pattern of brain involvement. Noteworthy, one-third of our patients revealed co-existent demyelination in varied areas across the brain (including the brainstem) in addition to the spinal cord. Laboratory parameters revealed elevated CSF protein and lymphocytic pleocytosis among reported cases of acute myelitis. These observations are also important from the perspective of a clinical neurologist. Concerning the prognosis, most of the patients came up with a complete disease resolution and the mortality rate was low (<10%). Noteworthy, microbes including *Mycobacterial pneumoniae, Epstein Barr Virus* (EBV), cytomegalovirus (CMV), rhinovirus, and measles are implicated in post infectious acute myelitis [31-33]. Reports have already suggested severe cytokine storm with significant elevation of IL-6, IL-7, IL-8, IL-9, TNF-alpha, IFN-gamma cytokines, and depletion of CD8^+^T_c_ cells and NK cells; i.e. lymphocytopenia [34].This event raises a possible hypothesis that the infectious organisms are targeted by the immune system which may also invade the CNS tissue including the spine due to structural similarities of microbial components and neuronal receptors. Different strains of coronaviruses such as HCoV-OC43 and MERS-CoV have been found to initiate several immunopathogenic responses which further cause the progression of demyelinating events in the central nervous system (CNS) [35]. The presence of HCoV RNA was found in the CNS of patients with demyelinating disorders, which suggests a possible association between coronaviruses and demyelination [36]. In a previous study, nucleic acids of HCoV-229E have been detected in the CNS tissue in four out of eleven multiple sclerosis (MS) patients, which suggest neurotropism of this species of coronavirus [37]. Moreover, coronavirus-like particles under electron microscopy were found from autopsies of two MS patients [38]. Using *in situ* hybridization technique with cloned coronavirus cDNA probes, Murray *et.al* detected coronavirus RNA in the demyelinating plaques in twelve out of twenty-two MS patients. Significant amounts of coronavirus antigen and RNA were observed in active demyelinating plaques from two patients with rapidly progressive MS [39]. A 3-year-old girl who was admitted with lower respiratory tract infection and acute flaccid paralysis showed co-infection with HCoV-229E and HCoV-OC43, which was detected by real-time PCR analysis of nasal swab samples [40]. Further, a 15-year old boy, who presented with acute disseminated encephalomyelitis (ADEM), tested positive for HCoV-OC43 in cerebrospinal fluid (CSF) using RT-PCR [41]. In a series of 4 patients affected with neurological complications due to MERS-CoV, one of them met a few criteria for diagnosis of the ADEM [42]. Furthermore, RNA recombination demonstrated that the S gene of the coronavirus mouse hepatitis virus (MHV) is related to certain molecular aspects of demyelination, which indicates the potential role of viral envelope S glycoproteins in autoimmune-induced demyelination [43].According to Kim *et al*, an experimental strain of CoVs JHM, even in the absences of T and B cells, developed an autoimmune demyelinating disorder in a mouse model suggesting that the formation of anti-JHM antibodies is sufficient to cause the demyelination. They also showed a decrement of such anti-JHM antibodies mediated demyelination by 90% and 76% in F_c_R_r_^-/-^ and complement depleting agents like cobra venom respectively. Overall observation suggests that direct autoimmune antibody formation against JHM structural protein is enough to start the cascade of demyelination [44]. Mutations in the spike glycoprotein of human coronavirus OC43 (HCoV-OC43) modulated the disease from chronic encephalitis to flaccid paralysis and demyelination in BALB/c mice [45]. Taken together, it seems that viral-mediated demyelination is very much evidential and concerning in times of a pandemic. Several cases of viral encephalitis have been found as another presentation of CNS involvement in context to acute COVID-19 indication. Interestingly, a few case reports, included in our study have also revealed the presence of SARS-CoV-2 RNA in CSF and brain MRI - findings consistent with meningoencephalitis, along with a simultaneous association of post or para-infectious acute myelitis. Wu *et al* hypothesized that SARS-CoV-2 may induce neuronal injury via hypoxic and immune mediated pathways. The presence of demyelination, as well as SARS-CoV-2 viral particles and genome sequence in the brain, has also been detected in autopsy studies [46, 47]. Besides, SARS-CoV-2 binds strongly with ACE2 receptors that have been distributed in kidney, lung, CNS, skeletal muscle, and the visceral tissues [ref-https://v15.proteinatlas.org/ENSG00000130234-ACE2/tissue]. Viral replication and subsequent turnover rate of ACE2 activation in CNS including spinal tissue may trigger systemic inflammatory response resulting in the increased permeability of blood brain barrier (BBB) and immune-mediated inflammation of the CNS [48]. Several other studies demonstrated that pathogenesis of severe viral infections is closely associated with the development of viral-induced severe inflammatory response syndrome (SIRS) like immune disorders [49] for SARS-CoV-2 infections. The pro-inflammatory state induced by cytokine storm mainly sustained by IL-6, IL-1, TNF-alpha may be the responsible for the activation of glial cells, which may also trigger the onset of demyelination [50].A possible hypothesis that comes up through such findings is the production of autoantibodies against glial cells triggered by the viral infection as a para or post-infectious phenomenon. Considering case reports and case series throughout this pandemic reporting multiple incidences of GBS [51] or GBS relate disorders (eg.-AMAN, AMSAN, and AIDP) [52] in COVID-19 along with profound documentation of viral encephalitis and encephalopathy; this systemic review elaborates unfathomable possibilities behind the onset of spinal cord demyelinating disorders, which needs to be addressed so that neurologists can be aware of the plethora of autoimmune neurologic complications involving CNS and can promptly be recognized and treated to minimize permanent neurologic disabilities and subsequent disease burden.

There are some implicit limitations in the present endeavor. Given the notable asymmetry between the total number of affected cases and reported cases of infection-related myelitis, it can be assumed that cases are presently under-reported which may be due to several reasons. Therefore, the present systematic review is based on a small number of cases even after an extensive search of available literature, both peer-reviewed, and pre-print. Also, several of the available reports do not describe the timeline of events in an organized manner making interpretation difficult. Laboratory features have also not been mentioned in detail in a few of the cases. In addition, there is considerable heterogeneity in the available data that may be considered a hindrance in advanced analysis. Despite these shortcomings, the present organized review will act as a preliminary guide for clinicians while dealing with suspected spinal cord demyelination in the context of SARS-CoV-2 infection.

## Data Availability

The data for this research is freely available online.

## Acknowledgement

We are sincerely thankful to Rohan Sarkhel (Department of Computer Science and Engineering, Maulana Abul Kalam Azad University, India) for his assistance in the literature search and preparing the illustrations for this paper.

